# Change in force profile of the hardstyle kettlebell swing in older adults is small following 16 weeks of training and may not be required to improve physical function: findings from the BELL trial

**DOI:** 10.1101/2021.08.27.21262528

**Authors:** Neil J. Meigh, Justin W.L. Keogh, Evelyne N. Rathbone, Wayne A. Hing

## Abstract

**Background:** Hardstyle kettlebell training is characterised by the ballistic two-handed kettlebell swing with outcomes believed to be strongly influenced by swing proficiency. This study examines the effect of four months hardstyle kettlebell training on the force profile of the two-handed kettlebell swing, and peak ground reaction force during a kettlebell deadlift in older adults. These data will help inform healthcare providers and coaches about the use and prescription of kettlebell exercises with older adults.

**Methods:** Five males and five females <70 years of age who participated in the BELL trial were recruited. Two-handed hardstyle swings were performed with 8-16 kg, and deadlifts with 8-32 kg. Ground reaction force (GRF) was obtained from a floor-mounted force platform. Force-time curves (FTCs), peak force, forward force relative to vertical force, rate of force development (RFD), and swing cadence were investigated. Results were compared with the same data variables collected from the participants in an exploratory pre-intervention study, conducted approximately seven months before the present study. Participants completed approximately 90 kettlebell training sessions during a four-month training intervention.

**Results:** Participants used kettlebells to perform 3779 ± 802 swings, 923 ± 251 cleans, 825 ± 309 snatches and 744 ± 178 deadlifts during group-training sessions. Peak ground reaction force during kettlebell swings did not significantly change with any kettlebell weight. There was a significant 3% increase in the magnitude of forward force during 8 kg swings, and a significant 3% decrease in forward force during 16 kg swings. There were large significant improvements in swing cadence with a mean increase of three swings per minute and a small non-significant increase in RFD. Change in kettlebell swing force-time curve profiles were small. Change in peak ground reaction force during deadlifts were moderate to large. All participants increased in grip strength following training, with the magnitude of change greater than the minimum clinically important difference for seven participants. All participants had significant increases in multiple secondary outcomes.

**Conclusion:** Group-based and online kettlebell training is likely to be ineffective for improving the force profile of the hardstyle kettlebell swing in older adults. Insufficiently active older adults engaged in high-volume kettlebell training performed 3-5 times weekly, can however expect to see clinically meaningful improvements in health-related physical fitness irrespective of swing proficiency, and have increased confidence with heavy lifting tasks. Results of this study suggest that beyond safe and competent performance, striving to optimise hardstyle swing technique may provide no additional benefit to clinical outcomes in older adults.

## Introduction

Hardstyle kettlebell training is believed to improve health-related physical fitness, particularly lower limb muscular power. Data from force-time curves show distinct differences between a novice and expert swing profile, with large effect size difference in ground reaction force ^(1)^. This suggests that technique proficiency significantly alters the force profile, but are improvements in technique and force profile required for the exercise to be effective? In the absence of motion analysis and kinematic differences which have previously been described ^(2)^, investigations of force profile demand suggests that a FTC profile from a force platform can be used as a fair proxy for determining the proficiency of a hardstyle swing ^(1, 3)^. If force profiles favorably change with improved swing proficiency and force profiles directly influence the adaptations to exercise, it could be hypothesised that improving hardstyle swing technique might enhance outcomes. Such improvements may be seen in athletic performance, the effects of therapeutic exercise in rehabilitation, or the management of adverse symptoms associated with conditions influenced by mechanical loading, such as knee osteoarthritis. Results from the BELL trial show large clinically meaningful increases in grip strength and significant improvements in cardiovascular capacity, muscular strength and endurance, functional capacity, and body composition in older adults, following a high-volume program of hardstyle kettlebell training ^(4)^. Participants trained five days a week accruing a mean training load exceeding 100,000 kg per person during the first 12 weeks of group training sessions. Results from the BELL trial also show improved physical function and capacity, and significant reductions in pain and disability associated with persistent low back and knee osteoarthritis ^(5)^. Significant increases in appendicular skeletal muscle mass, up to 2 kg, increased the skeletal muscle index in one female above the threshold for sarcopenia, and clinically significant increases in bone mineral density were observed in two participants with osteoporosis, sufficient for the status of one participant to advance from osteoporosis to osteopenia ^(6)^. It was previously proposed that knowledge of GRF during a hardstyle swing would assist a healthcare provider’s prescription of the hardstyle kettlebell swing as a therapeutic exercise, with high GRF combined with limited knee flexion and no impact loading, hypothesised to reduce knee pain in those with osteoarthritis ^(3)^. While the mediators and moderators of change remain unknown, hardstyle training principles suggest that technique would be a significant contributor, with the force profile and FTC profile of the participant’s swings expected to improve following four months of guided training. This is a longitudinal replication of a previous acute profiling study (3) using the same participants to perform kettlebell swings and kettlebell deadlifts under the same assessment conditions. The aim of this study is to report the change in force profile of a two-handed hardstyle kettlebell swing and kettlebell deadlift in older adults following 16-weeks of training. Findings are presented with outcomes from the BELL trial and discussed in the context of observing change in technique during six weeks of face-to-face coaching.

## Materials & Methods

### Participants

Five males and five females <70 years of age who participated in the BELL trial intervention took part in the study. Participants were free from injury with no health or medical conditions considered to be high risk. Participants were aware of the study aims, protocols, and potential risks, and gave their informed consent. Ethical approval for this study was granted by Bond University human research ethics committee (NM03279) which was subsumed within the larger BELL trial project (ACTRN12619001177145).

### Procedure

Participants performed kettlebell swings and deadlifts on a force flatform in October 2019, prior to their enrollment in the BELL. Results of the initial exploratory profile have been published elsewhere ^(3)^. Following a 3-month period of controlled activity, the BELL trial intervention commenced in February 2020. The participants did not use kettlebells between October 2019 and February 2020. Following the four-month BELL trial intervention, participants repeated the same swings and deadlifts as previously described ^(3)^. Participants were given the opportunity to perform a warm-up of their choice and encouraged to perform swings with an 8 kg kettlebell prior to the commencement of data collection. Participants received the same basic instruction about technique that they had received throughout the intervention period, and no additional coaching or correction was provided. Force USA competition kettlebells from 8-32 kg of standardised dimensions were used. Swings were performed with 8, 10, 12, 14 and 16 kg kettlebells. Deadlifts were performed with 8, 16, 24 and 32 kg kettlebells.

### Protocol

Data were collected from the University biomechanics laboratory with participants attending a single session of less than one hour in duration. The protocol was consistent with previous investigations ^(1, 3)^. Two-handed kettlebell swings to chest-height were performed on a floor-mounted force flatform (AMTI, Watertown, NY, USA) recording GRF at 1000 Hz using NetForce software (AMTI, USA). Participant body mass was captured by the force plate from a period of quiet standing. Tri-plantar force variables were obtained from the floor-mounted force platform. To gain some insight into how the force profile of the kettlebell exercises may have changed over the course of the training program, peak GRF, dynamic RFD and swing cadence were calculated. Participants performed a single set of 12 repetitions with each kettlebell, with the middle 10 repetitions used for analysis. A custom program (Microsoft Excel, Version 2012) was used to calculate peak force during each swing cycle of the set, with values manually assessed and verified against the corresponding FTC. To obtain peak net force, system weight (body mass + kettlebell weight) was subtracted from the square root of squared and summed data:

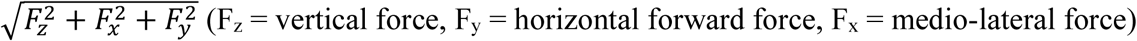

The back or bottom position of the swing was used as the start of each swing cycle. Dynamic RFD (N.s^-1^) during hip extension (propulsion) was calculated as the change in resultant GRF divided by elapsed time and normalised to body mass (N. s^-1^.kg^-1^), with this reported as the mean of 10 swings. Cadence in swings per minute (SPM) was calculated from the average time between peak force of hip extension in each swing cycle. Peak force was reported as resultant force unless stated otherwise.

### Statistical analyses

Descriptive statistics were reported for normally distributed continuous variables, with this data presented as mean (SD). Normality was verified using histograms, normal Q-Q plots, boxplots to support the Shapiro-Wilk test. Where the assumptions for the paired *t*-test were violated, the Wilcoxon signed rank test was used. Effect size was calculated using Lenhard and Lenhard ^(7)^, reported as *g*_Hedges_ and quantified as trivial, small, moderate, large, very large, and extremely large where effect size < 0.20, 0.20 - 0.59, 0.60 - 1.19, 1.20 - 1.99, 2.0 - 3.99 and ≥4.0 respectively ^(8)^. Statistical analyses were performed using SPSS (version 26.0; SPSS Inc., Chicago, IL, USA), and *p* < 0.05 was used to indicate statistical significance.

## Results

Participant demographics are presented in Table 1.

**Table 1.**
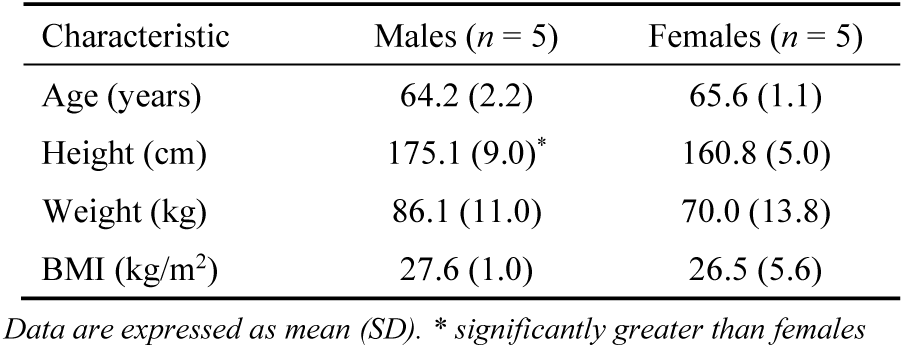
Participant characteristics (*N* = 10)

| Characteristic | Males (*n* = 5) | Females (*n* = 5) |
| --- | --- | --- |
| Age (years) | 64.2 (2.2) | 65.6 (1.1) |
| Height (cm) | 175.1 (9.0)* | 160.8 (5.0) |
| Weight (kg) | 86.1 (11.0) | 70.0 (13.8) |
| BMI (kg/m<sup>2</sup>) | 27.6 (1.0) | 26.5 (5.6) |

*Data are expressed as mean (SD). \* significantly greater than females*

Mean changes in force profile pre- to post-training are shown in Table 2. Resultant peak force during kettlebell swings did not significantly change with any kettlebell weight. There was a significant increase in the magnitude of forward force from 3% to 6% during swings with 8 kg, but a significant decrease in forward force from 7% to 4% with the 16 kg kettlebell, and no significant change during swings with 10, 12 or 14 kg. There were large significant improvements in swing cadence with a mean increase of 3 SPM. A small increase in the RFD during swings with 12 kg was not statistically significant. Moderate to large increases in resultant peak force during deadlifts were significant for 8 – 24 kg but not statistically significant with 32 kg.

**Table 2.** Pre- and post-training force profiles for the kettlebell swing and deadlift.

| Measure | Pre-test<br>Mean (SD) | Post-test<br>Mean (SD) | MD | 95% CI | *p*-value | *g*<sub>Hedges</sub> |
| --- | --- | --- | --- | --- | --- | --- |
| Swing – resultant peak force (N.kg) |  |  |  |  |  |  |
| 8 kg | 3.4 (1.1) | 3.9 (0.9) | 0.5 | -0.2, 1.2 | 0.137 | 0.44 |
| 10 kg | 3.9 (1.2) | 3.9 (1.0) | 0.0 | -0.8, 0.7 | 0.948 | -0.02 |
| 12 kg | 4.1 (0.8) | 4.1 (1.1) | 0.1 | -0.6, 0.7 | 0.844 | 0.05 |
| 14 kg | 4.3 (1.0) | 4.4 (1.4) | 0.1 | -1.2, 1.3 | 0.814 | 0.08 |
| 16 kg | 4.3 (1.3) | 4.5 (1.4) | 0.1 | -1.3, 1.5 | 0.797 | 0.09 |
| Swing – cadence (SPM) |  |  |  |  |  |  |
| 8 kg | 35.7 (2.1) | 39.2 (2.3) | 3.6 | 1.7, 5.4 | 0.002 | 1.45 |
| 10 kg | 35.6 (2.1) | 38.4 (2.3) | 2.8 | 0.7, 4.8 | 0.013 | 1.10 |
| 12 kg | 34.7 (2.4) | 38.6 (1.9) | 3.9 | 2.5, 5.2 | 0.005 | 1.65 |
| 14 kg | 35.4 (2.0) | 38.9 (1.8) | 3.5 | 1.5, 5.5 | 0.006 | 1.60 |
| 16 kg | 35.4 (1.9) | 37.5 (1.4) | 2.1 | 0.0, 4.2 | 0.053 | 0.96 |
| Swing – forward force (A/V) % |  |  |  |  |  |  |
| 8 kg | 3.2 (1.0) | 5.6 (2.9) | 2.4 | 0.04, 4.7 | 0.047 | 0.89 |
| 10 kg | 3.5 (1.6) | 5.0 (2.9) | 1.5 | -0.3, 3.3 | 0.097 | 0.53 |
| 12 kg | 4.1 (2.0) | 5.8 (3.9) | 1.7 | -0.4, 3.9 | 0.094 | 0.47 |
| 14 kg | 5.7 (3.1) | 5.4 (3.4) | -0.2 | -2.9, 2.4 | 0.820 | -0.01 |
| 16 kg | 6.6 (2.5) | 4.3 (3.3) | -2.3 | -4.5, -0.1 | 0.046 | -0.50 |
| Swing – rate of force development (N.s<sup>-1</sup>.kg.<sup>-1</sup>) |  |  |  |  |  |  |
| 12 kg | 17.0 (3.7) | 18.9 (4.9) | 1.9 | -1.0, 4.1 | 0.178 | 0.40 |
| Deadlift – resultant peak force (N.kg) |  |  |  |  |  |  |
| 8 kg | 2.6 (0.4) | 3.6 (0.6) | 1.0 | 0.5, 1.5 | 0.002 | 1.75 |
| 16 kg | 2.8 (0.6) | 3.6 (0.7) | 0.8 | 0.4, 1.3 | 0.003 | 1.19 |
| 24 kg | 2.9 (0.6) | 3.6 (1.0) | 0.7 | 0.1, 1.3 | 0.030 | 0.76 |
| 32 kg | 3.5 (0.4) | 4.1 (0.8) | 0.6 | -0.4, 1.6 | 0.133 | 0.71 |

MD = Mean Difference, SD = standard deviation, *g*<sub>Hedges</sub> = unbiased estimate of $\delta_{\text{Cohen}}$ , A/V = ratio of *anterior* (forward) horizontal to *vertical* ground reaction force, SPM = swings per minute.

Plots for paired data for resultant peak force, swing cadence, forward force, and RFD are shown in Figures 1 – 4, respectively, with each participant represented by connected data points shown in blue. Plots for resultant peak force during deadlifts are shown in Fig 5. Large positive and negative changes in resultant peak force during swings are evident at each kettlebell weight with negligible change in the mean for 10 – 16 kg (Fig 1). For swing cadence, 33 of 35 paired data sets show a positive change after training with moderate-to-large change in the mean at each weight (Fig 2).

**Fig. 1:**
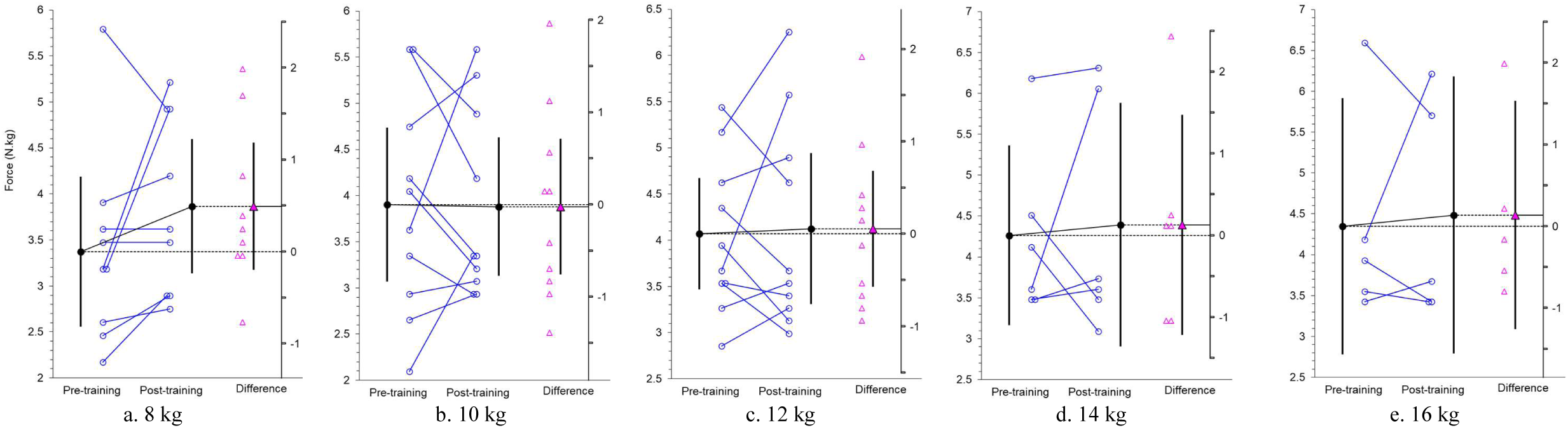
Change in resultant peak force during kettlebell swings (N.kg). • = mean, Δ = individual change, ▴ = mean change. Means displayed with 95% confidence interval (CI).

**Fig. 2:**
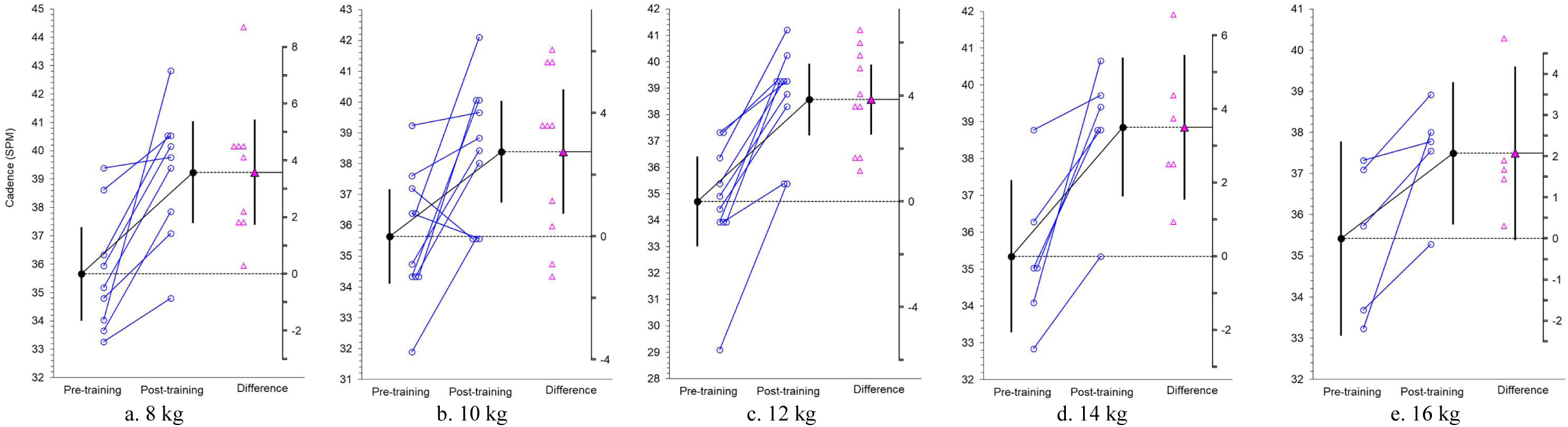
Change in kettlebell swing cadence (SPM = swings per minute). • = mean, Δ = individual change, ▴ = mean change. Means displayed with 95% CI.

**Fig. 3:**
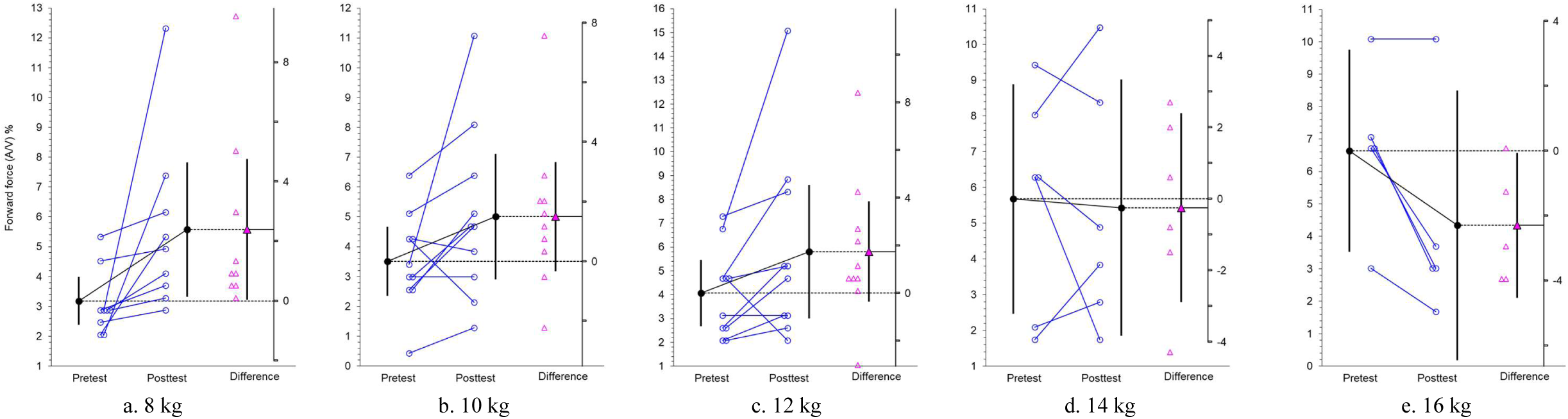
Change in magnitude of forward force relative to vertical force (%) during kettle bell swings (A/V = ratio of *anterior* (forward) horizontal to *vertical* GRF). • = mean, Δ = individual change, ▴ = mean change. Means displayed with 95% CI.

**Fig. 4:**
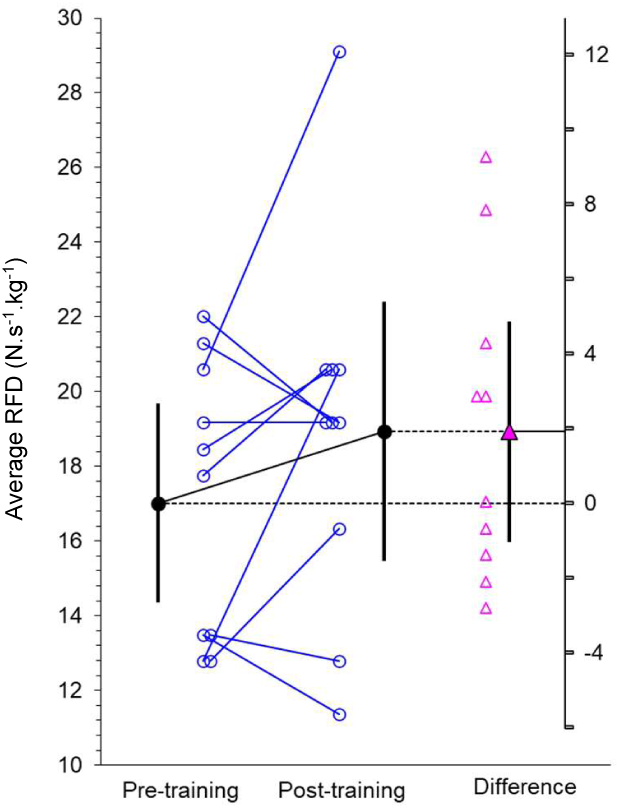
Change in rate of force development during a 12 kg kettlebell swing. • = mean, Δ = individual change, ▴ = mean change. Means displayed with 95% CI.

**Fig. 5:**
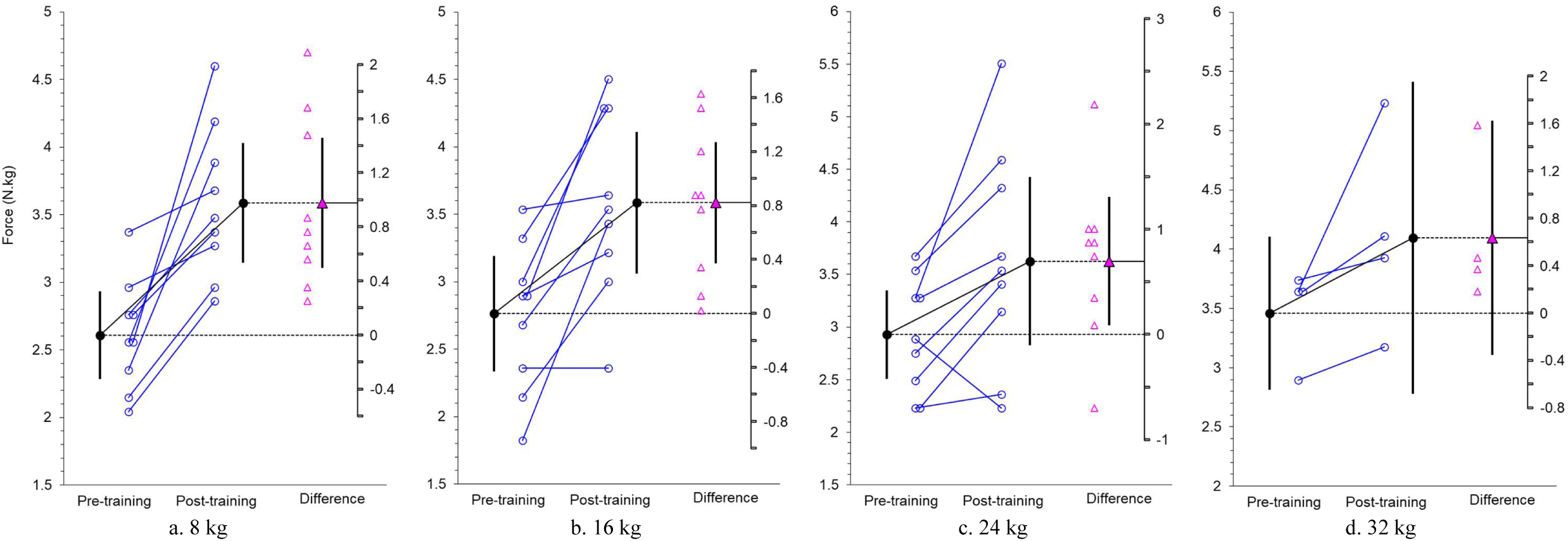
Change in resultant peak force during a kettlebell deadlift (N.kg). ♦ = mean, Δ = individual change, ▲ = mean change. Means displayed with 95% CI.

Paired data plots for forward force show a moderate mean increase for 8 kg swings, with an absolute difference less than 3%. There is a small increase in forward force for 10 and 12 kg swings, a trivial change for 14 kg, and a small decrease in forward force for 16 kg (Fig 3). There is a small increase in mean RFD for 12 kg swings of less than 2 N.s^-1^.kg.^-1^ which is statistically non-significant (Fig 4).

Paired data plots for kettlebell deadlifts show moderate-to-large increases in the mean peak ground reaction force for each weight, with a positive change in 29 or 33 paired data sets.

Change in force-time curve profiles were largely unremarkable (all participant FTC profiles are available as supplementary data). The FTC profile of a proficient hardstyle swing is characterised by a tall single narrow force peak closely followed by a second distinct force peak of smaller magnitude, with negligible change between 8 and 24 kg ^(1)^. In this study, some of the participants’ post-training FTC profiles had become more similar to a proficient swing i.e., suggestive of an improvement in technique (Fig. 6a). In some cases, however, the post-training FTC profile was less consistent with a proficient swing following training i.e., suggestive of a worsening of technique (Fig 6b). In most cases, the post-training FTC profile showed negligible change. An apparent worsening of an FTC profile, however, was not indicative of a negative change in force profile. For example, in one case where the FTC profile appeared to have worsened, RFD had increased by >30%. b. Change in force-time profile: *worsening*

**Fig. 6:**
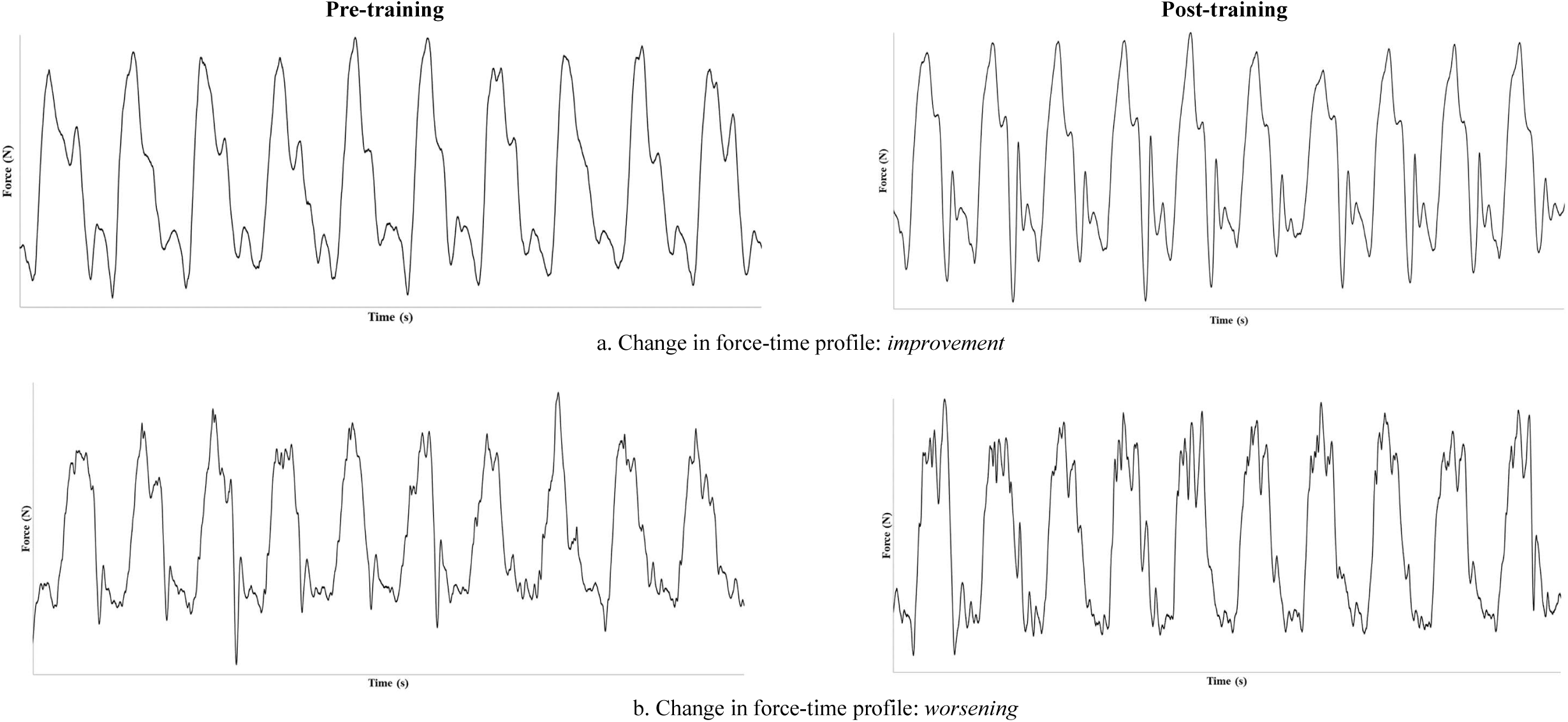
Change in kettlebell swing force-time curve profile pre- to post-training

Change in the participant’s primary and secondary outcomes of the BELL trial are shown in Tables 3 and 4, for females and males respectively. All participants in the present study had multiple significant improvements. During group training sessions (Monday, Wednesday, and Friday), participants in this study recorded 3779 ± 802 swings, 923 ± 251 cleans, 825 ± 309 snatches and 744 ± 178 deadlifts. Home-based training load volume was not recorded (Tue/Thur). Full details of the BELL trial intervention have been published elsewhere ^(4)^.

**Table 3.**
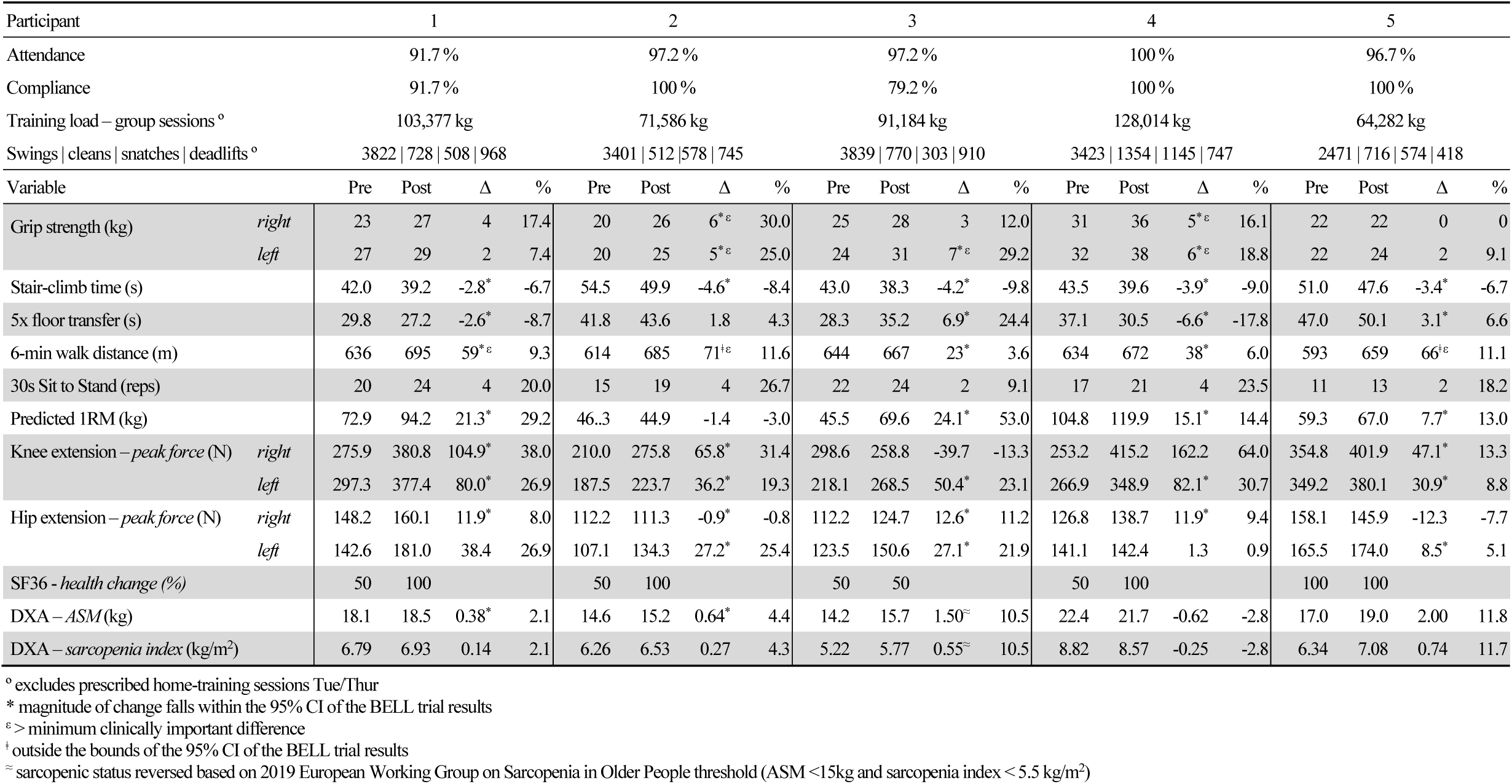
Individual change in outcome measures pre- to post-training - FEMALES

**Table 4.** Individual change in outcome measures pre- to post-training - MALES

| Participant |  | 6 |  |  |  | 7 |  |  |  | 8 |  |  |  | 9 |  |  |  | 10 |  |  |  |
| --- | --- | --- | --- | --- | --- | --- | --- | --- | --- | --- | --- | --- | --- | --- | --- | --- | --- | --- | --- | --- | --- |
| Attendance |  | 97.2 % |  |  |  | 97.2 % |  |  |  | 94.4 % |  |  |  | 100 % |  |  |  | 88.9 % |  |  |  |
| Compliance |  | 95.8 % |  |  |  | 95.8 % |  |  |  | 100 % |  |  |  | 100 % |  |  |  | 87.5 % |  |  |  |
| Training load – group sessions<sup>o</sup> |  | 165,588 kg |  |  |  | 107,088 kg |  |  |  | 120,740 kg |  |  |  | 150,750 kg |  |  |  | 101,022 kg |  |  |  |
| Swings \| cleans \| snatches \| deadlifts<sup>o</sup> |  | 5389 \| 977 \| 873 \| 991 |  |  |  | 3381 \| 934 \| 957 \| 594 |  |  |  | 4634 \| 1042 \| 1043 \| 767 |  |  |  | 4116 \| 1219 \| 1140 \| 662 |  |  |  | 3316 \| 981 \| 1131 \| 638 |  |  |  |
| Variable |  | Pre | Post | Δ | % | Pre | Post | Δ | % | Pre | Post | Δ | % | Pre | Post | Δ | % | Pre | Post | Δ | % |
| Grip strength (kg) | *right* | 45 | 49 | 4 | 8.9 | 42 | 52 | 10<sup>†</sup>ε | 23.8 | 32 | 41 | 9<sup>*ε</sup> | 28.1 | 30 | 45 | 15<sup>†</sup>ε | 50 | 35 | 44 | 9<sup>*ε</sup> | 25.7 |
|  | *left* | 55 | 59 | 4 | 7.3 | 43 | 44 | 1 | 2.3 | 27 | 34 | 7<sup>*ε</sup> | 25.9 | 26 | 40 | 14<sup>†</sup>ε | 53.8 | 35 | 40 | 5<sup>*ε</sup> | 14.3 |
| Stair-climb time (s) |  | 37.5 | 33.3 | -4.2* | -11.2 | 29.0 | 27.3 | -1.7* | -5.9 | 41.0 | 40.3 | -0.7* | -1.7 | 32.5 | 29.3 | -3.2* | -9.8 | 33.0 | 30.5 | -2.5* | -7.6 |
| 5x floor transfer (s) |  | 34.4 | 28.7 | -5.7* | -16.6 | 27.0 | 28.3 | 1.3 | 4.8 | 29.3 | 23.0 | -6.3* | -21.5 | 25.3 | 25.5 | 0.2 | 0.8 | 31.8 | 27.0 | -4.8* | -15.1 |
| 6-min walk distance (m) |  | 588 | 650 | 62<sup>*ε</sup> | 10.5 | 709 | 744 | 35* | 4.9 | 671 | 714 | 43* | 6.4 | 667 | 676 | 9 | 1.3 | 718 | 731 | 13 | 1.8 |
| 30s Sit to Stand (reps) |  | 12 | 16 | 4 | 33.3 | 18 | 24 | 6 | 33.3 | 10 | 24 | 14 | 140 | 14 | 14 | 0 | 0 | 16 | 19 | 3 | 18.8 |
| Predicted 1RM (kg) |  | 105.1 | 112.0 | 6.9* | 6.6 | 94.2 | 109.3 | 15.1* | 16.0 | 72.3 | 112.3 | 40.0 | 55.3 | 85.0 | 97.0 | 12.0* | 14.1 | 72.6 | 84.3 | 11.7* | 16.1 |
| Knee extension – peak force (N) | *right* | 368.7 | 384.8 | 16.1* | 4.4 | 337.8 | 535.9 | 198.1 | 58.6 | 316.7 | 394.7 | 78.0* | 24.6 | 339.2 | 332.6 | -6.6 | -1.9 | 476.5 | 402.1 | -74.3 | -15.6 |
|  | *left* | 290.4 | 451.6 | 161.2 | 55.5 | 324.6 | 456.7 | 132.1 | 40.7 | 374.6 | 360.1 | -14.4 | -3.9 | 334.9 | 360.1 | -14.4 | -3.9 | 491.1 | 452.7 | -38.5 | -7.8 |
| Hip extension – peak force (N) | *right* | 183.7 | 204.0 | 20.3* | 11.0 | 237.7 | 230.0 | -7.7 | -3.3 | 170.5 | 177.3 | 6.8* | 4.0 | 160.6 | 192.4 | 31.9 | 19.8 | 232.2 | 276.3 | 44.1 | 19.0 |
|  | *left* | 216.6 | 187.4 | -29.2 | -13.5 | 201.6 | 245.5 | 44.0 | 21.8 | 184.0 | 195.0 | 11.1* | 6.0 | 184.9 | 208.6 | 23.7* | 12.8 | 244.8 | 272.8 | 28.0* | 11.4 |
| SF36 - health change |  | 50 | 75 |  |  | 50 | 50 |  |  | 50 | 75 |  |  | 75 | 100 |  |  | 50 | 50 |  |  |
| DXA – ASM (kg) |  | 29.0 | 30.3 | 1.29 | 4.4 | 26.0 | 27.3 | 1.27 | 4.9 | 18.6 | 19.1 | 0.52* | 2.8 | 22.6 | 23.4 | 0.79* | 3.5 | 22.2 | 22.4 | 0.19* | 0.8 |
| DXA – sarcopenia index (kg/m2) |  | 8.47 | 8.85 | 0.38 | 4.5 | 7.91 | 8.29 | 0.38 | 4.8 | 7.09 | 7.29 | 0.20 | 2.8 | 7.57 | 7.83 | 0.26 | 3.4 | 7.34 | 7.41 | 0.07 | 1.0 |

<sup>o</sup> excludes prescribed home-training sessions Tue/Thur
\* magnitude of change falls within the 95% CI of the BELL trial results
ε > minimum clinically important difference
† outside the bounds of the 95% CI of the BELL trial results

## Discussion

The findings of this study show that 16-weeks of kettlebell training had minimal effect on improving the force profile and FTC profile of a hardstyle kettlebell swing in novice older adults. Change in outcomes from the BELL trial, however, show that significant improvements in health-related physical fitness were achieved without such improvements. These findings indicate that, in contrast to the guidelines for prescribing resistance training for older adults ^(9)^, there may be cases when resistance training can still provide clinically important benefits even when technique is not optimised, and without increasing risk of injury. Acquiring formal training in hardstyle practice is still recommended for people training with kettlebells, but healthcare providers without such training might feel encouraged that older adults are likely to see benefit from engaging in kettlebell training, irrespective of their technical proficiency and the apparently limited capacity for swing kinetics to be improved within a group-exercise setting.

Our proposition that kettlebell swings could be an effective therapeutic exercise for the management of symptomatic knee osteoarthritis ^(3, 10)^ appears to be supported by results of the BELL trial ^(4, 5)^ however, the results of this study indicate that symptoms can be improved independent of a change in peak GRF during swings. Over 60% of the total treatment effect of non-nonsurgical interventions for knee osteoarthritis have been attributed to contextual effects ^(11)^, although this may also not be true of exercise-based interventions. There is also evidence that personality traits of the healthcare provider, specifically when low in neuroticism, might also significantly reduce the severity of complaints associated with chronic diseases such as arthritis ^(12)^. This is consistent with the results of the BELL trial, in which the instructor’s personality (low in neuroticism and volatility ^(13)^) was reported by the participants to have been a major factor in their engagement ^(5)^. There is evidently more to the effective management of osteoarthritic knee pain than changing kinetics. Kettlebell swings may be a helpful therapeutic intervention for managing knee osteoarthritis, but they are unlikely to be a panacea when used in isolation.

The ballistic hardstyle swing is believed to increase hip extension power ^(14)^. In this study, the 11.2% increase in RFD during swings was similar to the magnitude of change in isometric hip extension RFD reported in the BELL trial ^(4)^. It remains a feasible association although causation cannot be established from these data. The absence of a significant change in lower limb power and only small changes in hip and knee extension RFD in the BELL trial, suggest that kettlebell training may not be the most effective means of improving these qualities among older adults for whom this measure is clinically important e.g., individuals at higher risk of falls. Analysis of paired data in this study, shows large improvements for some participants in each of the kinetic measures, but negative changes for other participants. Further investigation is required to identify the factors which lead to someone being a responder or non-responder to kettlebell training. The association between swings, lower limb power, and falls risk, remains a valid hypothesis and should be tested under fair conditions.

It was encouraging that mean post-training swing cadence was approaching the idealized 40 SPM reported in hardstyle studies ^(10)^ and which is typically seen in practice. An increase of almost 4 SPM with the 12 kg kettlebell is sufficient to explain the small 2 N.s^-1^.kg^-1^ increase in RFD; however, it was unexpected that peak GRF did not also increase. In proficient swings there is a very strong positive correlation with kettlebell weight and peak GRF ^(1)^, thus GRF had been expected to increase following training. A distinct movement strategy previously reported in novice older adults of actively flexing the shoulders to ‘lift’ the bell during its’ upward trajectory ^(3)^, appears to have persisted throughout the intervention period for some of the participants in this study. It was expected that this too would have diminished or been eliminated with training. Although it is easier to swing a heavier kettlebell upwards than to lift it, the potential benefit for older adults performing swings with heavier kettlebells e.g., > 32 kg, remains unclear. Additionally, the potential benefit for younger adults swinging very heavy kettlebells e.g., > 48 kg, is also unclear. These results suggest that 3-4 months of group-based and online training is likely insufficient to improve the hardstyle swing proficiency in novice older adults, and a more tailored approach would be required for significant improvements to occur.

The hardstyle kettlebell swing is a ballistic exercise of the lower limbs. Force acting on the kettlebell is transmitted through a stiffened trunk and straight but relaxed upper limbs. It could be argued that greater upper body activity to ‘lift’ the bell, the factors which hardstyle training seeks to reduce or eliminate, could be beneficial in some cases. Older adults need to be able to bend, carry, and lift objects during activities of daily living (ADLs). Typically, these tasks, such as lifting a heavy basket of washing from the floor, would not be performed rapidly, thus it could be argued that the ideal hardstyle swing may provide less carryover to ADLs for older adults. In this instance, emphasising a ‘lifting’ pattern during a kettlebell swing may provide greater carryover to ADLs. What McGill and Marshall ^(15)^ describe as “street wisdom”, suggests that a large proportion of ground reaction force during a hardstyle swing is said to be in the forward horizontal direction. This belief is inconsistent with the data ^(1, 3, 16)^. It was surprising that the magnitude of forward force only significantly increased with the lightest kettlebell and decreased with the 16 kg kettlebell. It remains unclear whether there is any benefit in trying to increase the magnitude of horizontal force during a hardstyle swing.

It is likely that the participant’s swing proficiency could have been improved under different training conditions. Due to the limitations of a group-based design and the large variation in physical capacity within the participants of the BELL trial, accumulation of training load volume was prioritised ahead of technique. As a result, participants received less instruction on technique than would typically be seen in hardstyle practice. Nonetheless, given the instruction that was provided, daily training, and the high number of repetitions performed over a four-month training period, noticeable improvements in FTC profiles had been expected. The FTC profile of a proficient hardstyle swing is characterised by a tall single narrow force peak closely followed by a second distinct force peak of smaller magnitude, with negligible change between 8 and 24 kg ^(1)^. Using FTC profiles as a proxy for hardstyle swing technique, it was surprising, although consistent with the kinetic data, that most participant’s FTC profiles did not show more of the characteristics of a profile swing profile. This provides further evidence that the potential effect or benefits of the hardstyle swing might be achievable without improvement in technique.

Progressive resistance training improves leg extensor strength with moderate to large effect in older adults ^(17)^. Contrary however to the claims that the hardstyle kettlebell swing increases lower limb power ^(14)^, a recent meta-regression shows that explosive movements may be no better than strength training for improving RFD in older adults ^(18)^. The reported effect of resistance training on improvements in RFD is 14.4 – 35.5%. Following the BELL trial intervention, change in hip extension RFD was 6.7 – 12.7%, and knee extension slightly higher at 17.4 – 19.1% ^(4)^, both findings below the expected mean of 26.7% from random effects modelling of previous studies ^(18)^. The trivial to small changes in RFD reported in the BELL trial did not however translate to a significant change in relative lower limb power measured during sit-to-stand performance, or absolute lower limb power expressed as vertical jump height ^(4)^. There was however a significant 23% increase in the number of 30-second sit-to-stand repetitions; a similar effect to that reported in a meta-analysis of jump training in older adults ^(19)^. Distinct kinematic differences between novice and expert hardstyle swing have previously been reported ^(1)^, thus proficiency must be considered when interpreting data from novices. As such, these findings are only weak evidence that the hardstyle swing has negligible or no effect on improving lower limb power.

Large significant increases in peak ground reaction force during deadlifts were unsurprising. Participants reported feeling stronger and more confident lifting heavy objects following training ^(5)^, which is sufficient to explain the observed change in this study. Increased confidence and decreased hesitancy in initiating movement could account for an increased rate of acceleration of the bell being lifted from the ground. A large recent meta-analysis of exercise training on handgrip strength in healthy community-dwelling older adults over 60 years of age, showed only a small pooled transfer effect of 0.28 (SMD) ^(20)^. This limited effect was explained by a lack of specificity and insufficient training volume (training sessions too infrequent and duration too short). The mean difference in the BELL trial was larger than all of the 24 studies included in the meta-analysis by Labott et al ^(20)^. The reported effect from meta-analysis of resistance training on improvements in muscle strength was 13.7 – 23.3% ^(18)^. In the BELL trial, the effect on 1RM was an improvement of 23.3% ^(4)^. These results show that kettlebell training has a considerably larger effect on grip strength than other exercise programs typically prescribed for older adults, and improvements in 1RM from kettlebell training can be expected to meet or exceed those of any other form of community-based resistance training program for older adults.

Results of the BELL trial show a strong positive correlation in external training load volume and arbitrary training units over time ^(4)^, with all participants recording a “very hard” or “maximal effort” sRPE on the final day of the intervention. Although training volume remained high, participants did not have access to heavy kettlebells after the transition to home-only training at the end of week six. In addition, the rate of supervision dropped from 60% to 26% due to COVID-19 restrictions which prevented continuation of face-to-face training. As supervision and intensity both significantly affect outcomes ^(21, 22)^, it is likely that greater improvements could have been achieved. Additionally, a group-exercise format was not an ideal fit for some of the participants with the highest and lowest physical capacity, with smaller class sizes recommended ^(5)^. Furthermore, individually tailored programs are also likely to facilitate better clinical outcomes in some cases ^(6)^. Group-based training provides psychosocial benefits and fosters high rates of engagement; thus, program design and delivery is an important consideration in meeting the diverse needs of older adults.

## Limitations

This study is not without limitations. Firstly, participants >70 years of age were excluded from the study. Results from a sample size of 5 females and 5 males therefore may not be an accurate representation of effects for all participants in the BELL trial. Secondly, participants are otherwise healthy older adults. Results should not be generalised to different populations without due consideration. Thirdly, rate of force development should be interpreted with caution. In the absence of concurrent video analysis, calculation of RFD from force-plate data is subject to interpretation and reduces reliability. Finally, ground reaction data from hardstyle swings cannot be generalised to the double knee-bend swing (kettlebell Sport) or overhead (American) swing which are kinematically different ^(23, 24)^. Given that differences in force profile during the hardstyle swing have previously been attributed to technique, these findings should not be used as evidence that technique is not important or does not influence outcomes, as the findings of this study might be explained by the group-based study design rather than the mode of exercise. For some populations, in some cases, it may be more beneficial for the individual to simply accrue training load instead of prioritising adopting all the tenants of hardstyle swing technique. A proficient swing can be determined from an FTC profile, but the utility of these in clinical practice is unclear and likely of little benefit to healthcare providers. Motion analysis, either by visual observation or by slow motion video capture, remains the best means for trainers and coaches to assess swing proficiency, and a helpful tool for providing instruction.

## Conclusion

In conclusion, moderate to high-intensity group-exercise using hardstyle kettlebell training can promote healthy aging in older adults, however these benefits appear not to be dependent upon an ideal swing technique. Negligible improvement in swing performance was observed following a four-month program of training five days a week. Insufficiently active older adults can attain the health benefits of kettlebell training despite their swing technique rather than because of it, with little evidence that striving to optimise technique will provide additional benefit for the outcomes which are clinically important or meaningful. Further research is warranted to investigate the effects of kettlebell training with older adults using single case experimental designs, to address individual needs and better understand the relationships between hardstyle techniques and clinical outcomes of interest.

## Supporting information

Supplementary data - change in FTC profiles

## Data Availability

Data will be made available in accordance with Bond University Research Data Management and Sharing Policy (TLR 5.12, Sections 9.3, 10.2. 10.5, 10.8). Research data and primary materials will be made openly available for use by other researchers and interested persons for further research after a reasonable period following completion of the research.

## Acknowledgements

The authors would like to acknowledge Mr Benjamin Hindle for his support in the calculation of rate of force development.

## Contributions

NM recruited the participants and conducted the study, curated and analysed the data, interpreted the results, conducted the formal analysis, and wrote the original draft. JK and WH supported with ongoing consultation. ER provided direction with statistical analysis. JK and WH reviewed and provided revisions to earlier versions of the manuscript. All authors read and approved the final manuscript.

## Ethics declarations

### Ethics approval and consent to participate

Ethical approval for this study was granted by Bond University human research ethics committee (NM03279) which was subsumed within the larger BELL trial project (ACTRN12619001177145). Participants were aware of the study aims, protocols, and potential risks, and gave their informed consent.

### Consent for publication

Not applicable

### Rights and permissions

Open Access This article is distributed under the terms of the Creative Commons Attribution 4.0 International License (http://creativecommons.org/licenses/by/4.0/), which permits unrestricted use, distribution, and reproduction in any medium, provided you give appropriate credit to the original author(s) and the source, provide a link to the Creative Commons license, and indicate if changes were made. The Creative Commons Public Domain Dedication waiver (http://creativecommons.org/publicdomain/zero/1.0/) applies to the data made available in this article, unless otherwise stated.

