## Supplementary data - change in FTC profiles for "Change in force profile of the hardstyle kettlebell swing in older adults is small following 16 weeks of training and may not be required to improve physical function: findings from the BELL trial"

**Supplementary data:** Change in force-time curve profiles before and after training intervention

Before intervention

After intervention

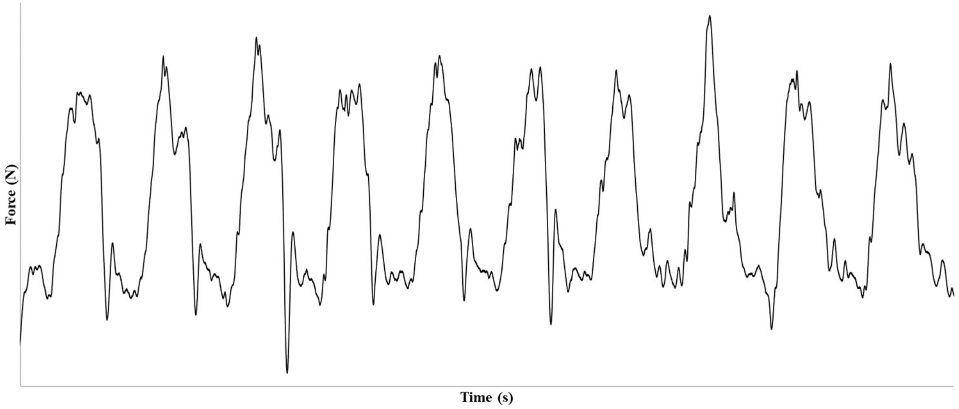


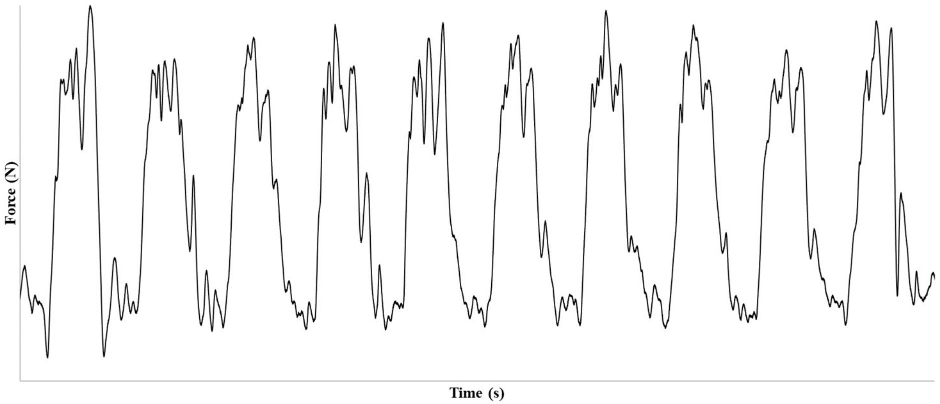


Participant 1

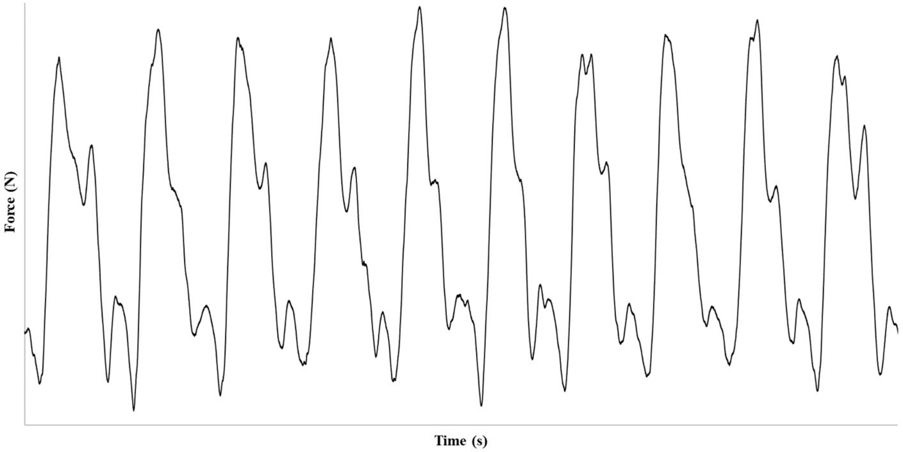


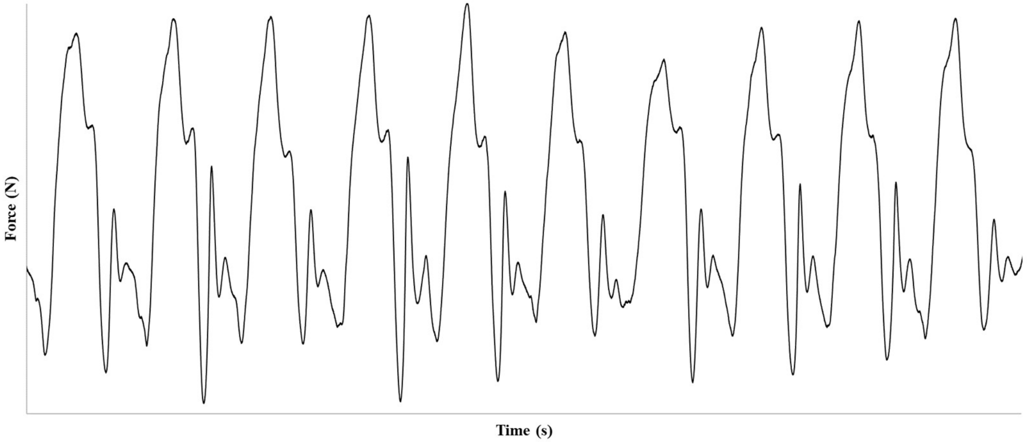


Participant 2

Before intervention | After intervention

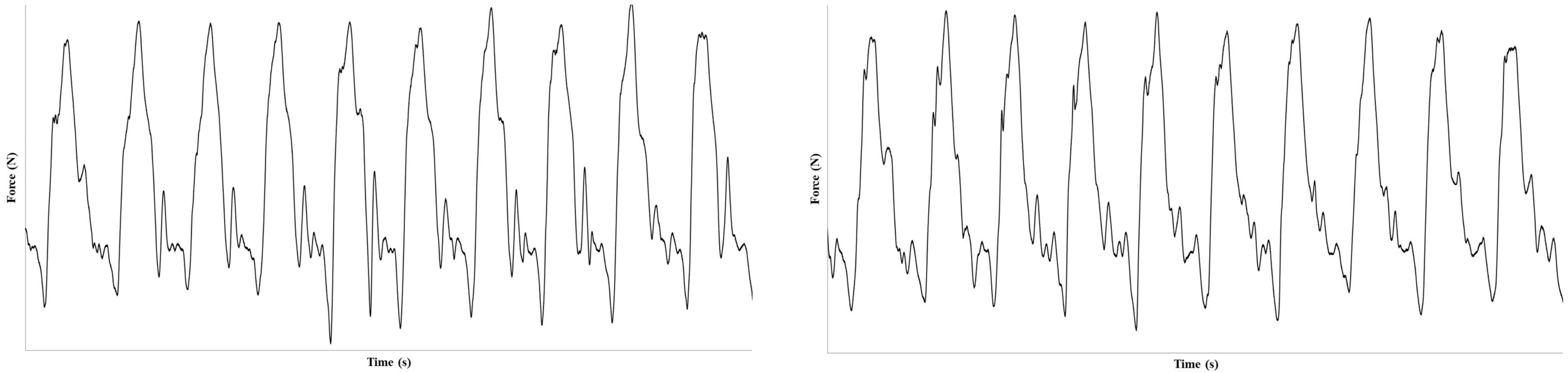


Participant 3

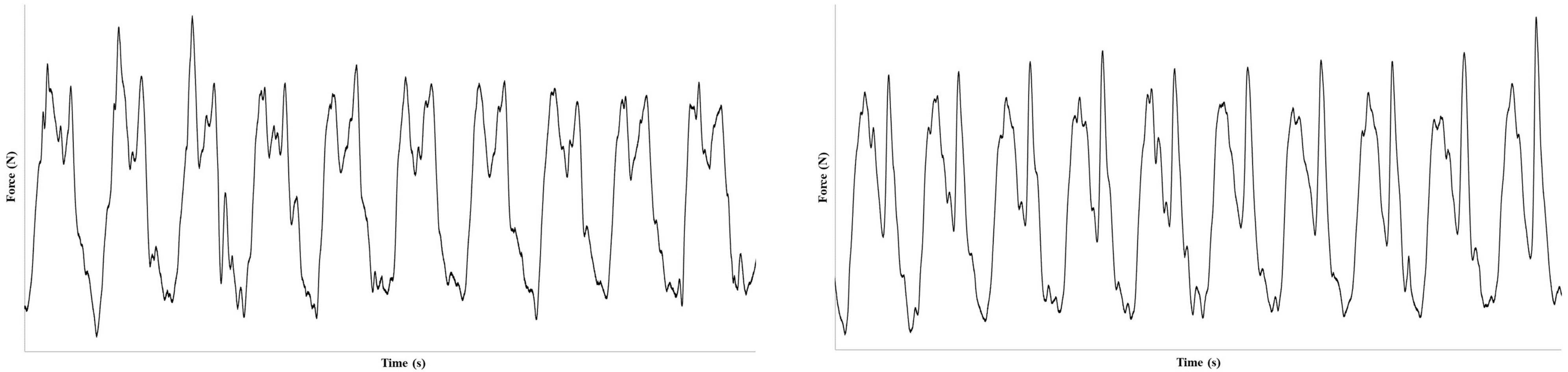


Participant 4

Before intervention

After intervention

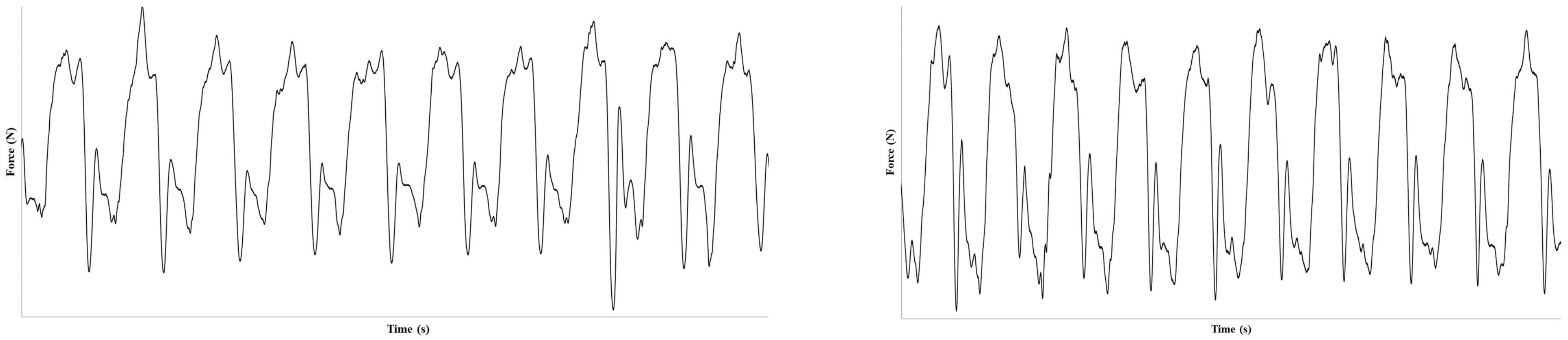


Participant 5

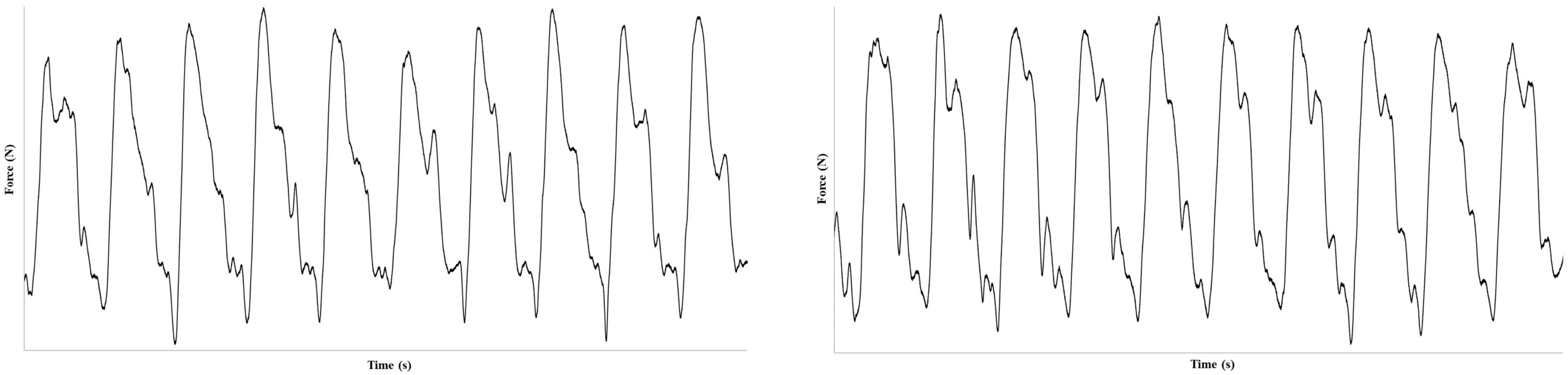


Participant 6

Before intervention

After intervention

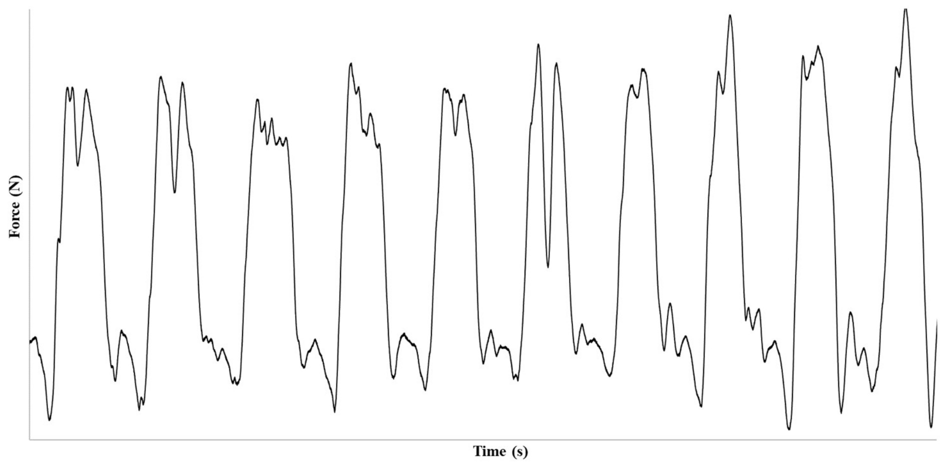


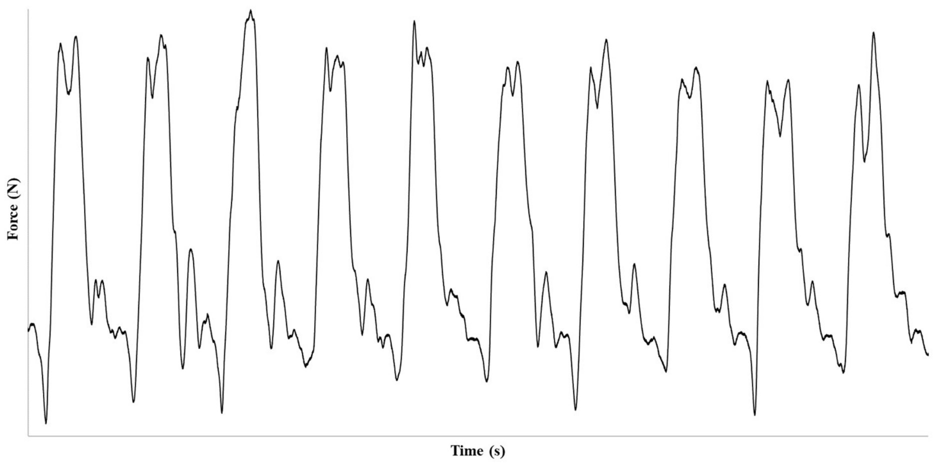


Participant 7

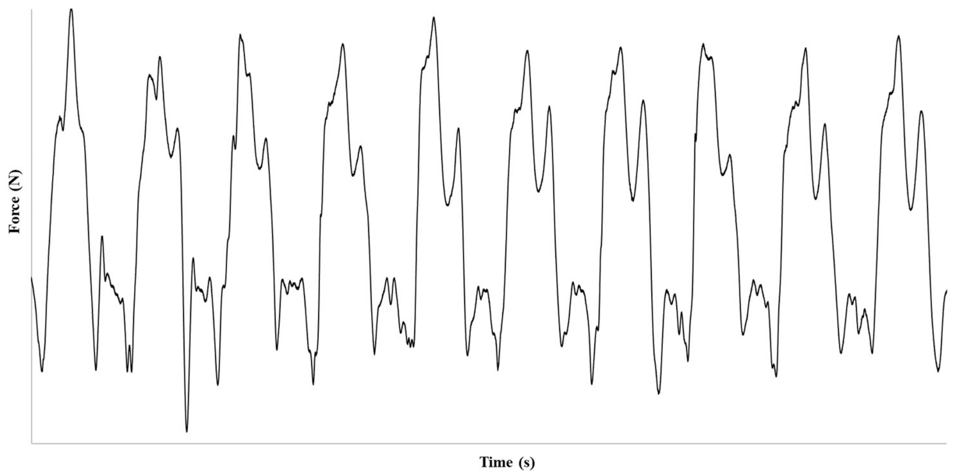


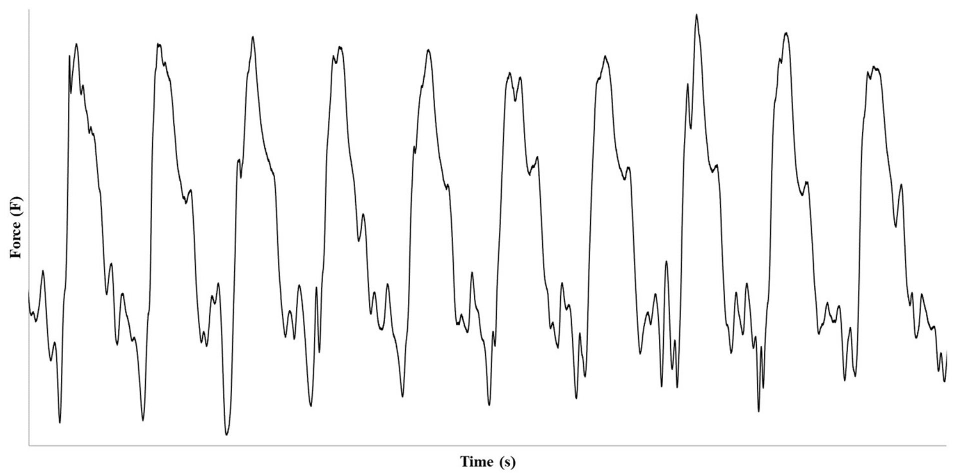


Participant 8

Before intervention

After intervention

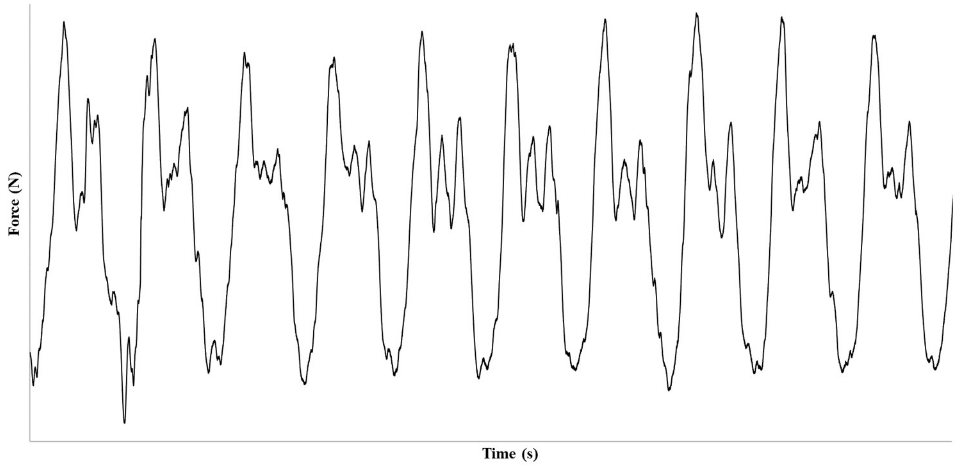


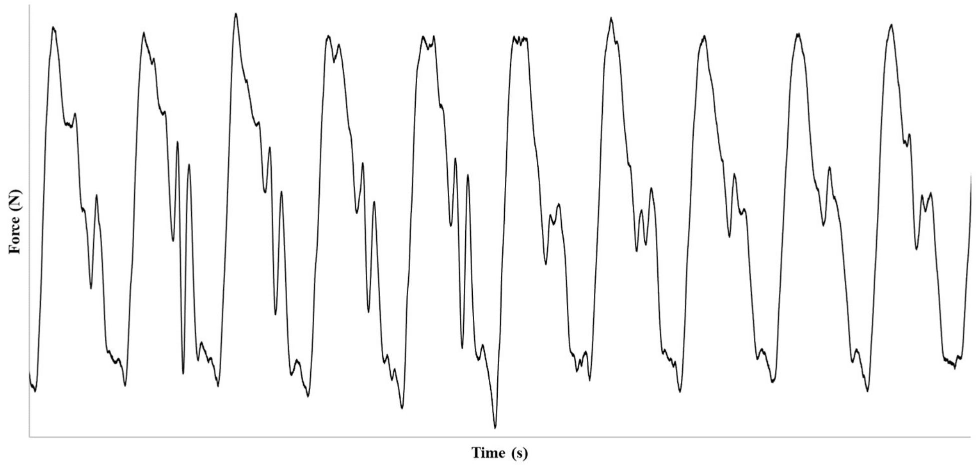


Participant 9

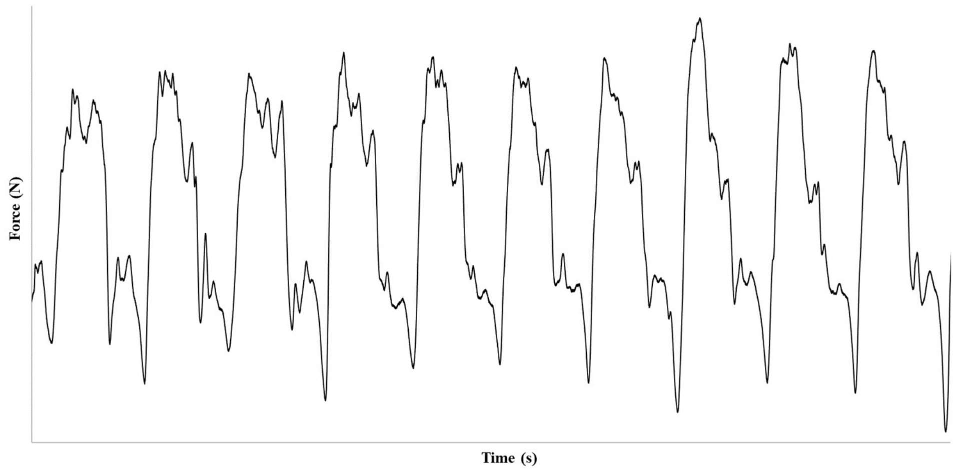


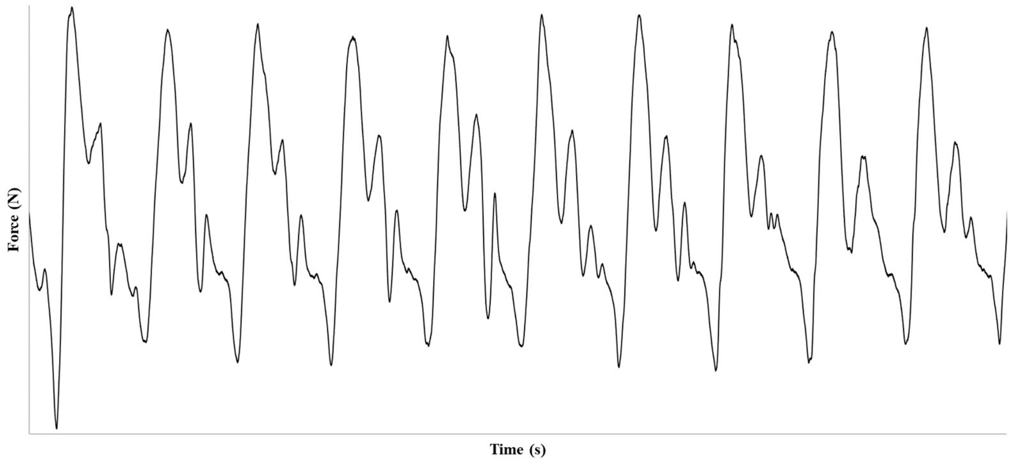


Participant 10
